# Development and Validation of a Web-Based Severe COVID-19 Risk Prediction Model

**DOI:** 10.1101/2020.07.16.20155739

**Authors:** Sang H. Woo, Arturo J. Rios-Diaz, Alan A. Kubey, Dianna R. Cheney-Peters, Lily L. Ackermann, Divya M. Chalikonda, Chantel M. Venkataraman, Joshua M. Riley, Michael Baram

**Author notes:** **Correspondence:** Sang Hoon Woo, M.D., Division of Hospital Medicine, Thomas Jefferson University Suite 701, 833 Chestnut St., Philadelphia, PA 19107. The authors have no conflicts of interest to disclose related to this manuscript. No funding was obtained for this manuscript. The content of this manuscript was not presented at any prior conferences.

## Abstract

**Background:** Coronavirus disease 2019 (COVID-19) carries high morbidity and mortality globally. Identification of patients at risk for clinical deterioration upon presentation would aid in triaging, prognostication, and allocation of resources and experimental treatments.

**Research Question:** Can we develop and validate a web-based risk prediction model for identification of patients who may develop severe COVID-19, defined as intensive care unit (ICU) admission, mechanical ventilation, and/or death?

**Methods:** This retrospective cohort study reviewed 415 patients admitted to a large urban academic medical center and community hospitals. Covariates included demographic, clinical, and laboratory data. The independent association of predictors with severe COVID-19 was determined using multivariable logistic regression. A derivation cohort (n=311, 75%) was used to develop the prediction models. The models were tested by a validation cohort (n=104, 25%).

**Results:** The median age was 66 years (Interquartile range [IQR] 54-77) and the majority were male (55%) and non-White (65.8%). The 14-day severe COVID-19 rate was 39.3%; 31.7% required ICU, 24.6% mechanical ventilation, and 21.2% died. Machine learning algorithms and clinical judgment were used to improve model performance and clinical utility, resulting in the selection of eight predictors: age, sex, dyspnea, diabetes mellitus, troponin, C-reactive protein, D-dimer, and aspartate aminotransferase. The discriminative ability was excellent for both the severe COVID-19 (training area under the curve [AUC]=0.82, validation AUC=0.82) and mortality (training AUC= 0.85, validation AUC=0.81) models. These models were incorporated into a mobile-friendly website.

**Interpretation:** This web-based risk prediction model can be used at the bedside for prediction of severe COVID-19 using data mostly available at the time of presentation.

## INTRODUCTION

COVID-19 is caused by the severe acute respiratory syndrome coronavirus 2 (SARS-CoV-2).^1^ The disease has spread rapidly throughout the world with more than 6 million confirmed cases and 372,035 deaths as of June 1st, 2020.^2^ Mortality in the United States b(U.S.) is over 100,000.^2,3^ COVID-19 is associated with a high fatality rate, roughly 6% worldwide with variation by country.^2,4–6^ The severity of COVID-19 illness varies from asymptomatic to severe disease that requires ICU admission.^7–10^ Patients with severe disease who are admitted to the ICU and require mechanical ventilation experience the highest mortality, reported as high as 53.4%.^9,11,12^ Part of COVID-19’s complexity is its variable time course and severity.(**e-Figure 1**) Therefore, early identification of patients at risk for progression to severe COVID-19 is paramount for accurate triage, determining appropriate diagnostic and treatment approaches, and resource allocation.

The progressing health crisis precipitated by this pandemic has been exacerbated by the lack of data on key clinical factors associated with severe presentation of disease. Emergent studies have reported several independent risk factors for the development of severe adverse outcomes among patients with COVID-19.^13,14^ In the U.S., available COVID-19 studies are descriptive in nature.^9,15,16^ A large study retrospectively analyzed 5,700 hospitalized patients from 12 hospitals in the New York City area. Comorbidities such as hypertension (57%), obesity (42%), and diabetes (34%) were prevalent in this cohort.^9^ For U.S. patient data, there are no effective point-of-care prediction models to identify COVID-19 patients at the highest risk for mortality. According to a recent systematic review, most prediction models available by the end of March 2020 were developed using patient data from China,^17^ and it is unclear if they are generalizable to a U.S. population. Furthermore, there are no effective prediction models specifically for identifying patients at risk of progressive disease that requires ICU admission and use of mechanical ventilation.

An effective COVID-19 risk prediction model has the potential to aid clinical guideline updates and guide scarce resource allocation, such as personal protective equipment, ventilators, hospital and ICU beds, extracorporeal membrane oxygenation, and potential treatments. It was reported that acute illness scores, such as the Sequential Organ Failure Assessment (SOFA) score were unlikely to predict critical care outcomes accurately.^18,19^ A risk stratification model could also help standardize portions of COVID-19 research by deterioration risk. Thus, we created a prognostic model to identify severe COVID-19 to be used on hospital admission. This model was developed and validated in an urban environment with a high prevalence of COVID-19. The cohort consists of diverse patients within a large academic medical center and four affiliated, community hospitals serving three adjacent states in the U.S..

## METHODS

### Study design, population

This retrospective cohort study included adult patients (>18 years) admitted to a large academic medical center and community hospitals with a diagnosis of COVID-19, defined as positive SARS-CoV-2 polymerase chain reaction (PCR). Patients were admitted between March 1st and April 30th, 2020. The study exclusion criteria were transfer from outside hospital with no access to the data, admission to labor and delivery, and admission for a primary surgical or trauma reason. Patients were grouped according to the severity of COVID-19. Non-severe COVID-19 was defined as requiring hospitalization but not meeting the definition of severe COVID-19 (defined as intensive care unit (ICU) admission, mechanical ventilation, and/or death).

### Data source and covariates

Demographics, clinical information, laboratory findings at the time of initial presentation, as well as outcome data were obtained through review of the electronic health record (EHR, Epic Systems Corporation, Verona, WI). Covariates included age, sex, self-reported race (White, Black, Asian, Hispanic), smoking status, body mass index, comorbidities (coronary artery disease [CAD], congestive heart failure [CHF], chronic obstructive pulmonary disease [COPD], interstitial lung disease [ILD], asthma, diabetes mellitus type 1 or 2, hypertension [defined by use of hypertension medications], end-stage renal disease [ESRD; defined by chronic dialysis]), chronic steroid use, history of cancer, dyspnea upon presentation [defined as subjective report of difficulty breathing, shortness of breath, or caretaker reporting they appear as such], white blood cell count (WBC), absolute lymphocyte count, hematocrit, platelet count, serum sodium, serum creatinine, C-reactive protein (CRP; 0-5mg/dL, 5-15mg/dL, >15mg/dL), elevated troponin (HsTroponin T > 19ng/L; Troponin I ≥ 0.04ng/mL), creatine kinase (CK), ferritin (0-300ng/mL, 300.1-1000ng/mL, >1000ng/mL), lactate, lactate dehydrogenase (LDH), aspartate aminotransferase (AST; 0-80 IU/L, >80 IU/L), D-dimer (ng/mL), and acute kidney injury (defined as increase in creatinine ≥ 0.3mg/dL from baseline) upon admission to the hospital. The data points extracted were verified by two independent reviewers, with consensus reached by a third independent reviewer if disagreement was noted. Information on race was limited to data available within the EHR system, which classifies Hispanic as a distinct category rather than under a separate category for ethnicity as defined by the U.S. Census Bureau.^20^

### Outcomes

The primary outcome was a composite measure defined by ICU admission(s), use of mechanical ventilation, and/or death within 14 days of hospitalization (referred to as “severe COVID-19”). These outcomes were also assessed independently. The secondary outcome was death within 14 days of hospital admission. When patients were discharged before the follow-up period (14 days), outcome assessment was carried through readmissions if records were available.

### Statistical analysis, model development and validation

Counts and frequencies were used to report categorical data. Means with standard deviation (SD) and medians with interquartile ranges (IQR) were used to report normally and non-normally distributed data, respectively. Chi-squared tests were used for comparison of categorical data. T-tests and Wilcoxon rank sum tests were used to compare continuous data as appropriate.

The eligible sample (n=415) was randomly split into a derivation group (75%; n=311) and a validation group (25%; n=104). The prediction risk model was created using the derivation sample. The derivation and validation groups were randomly selected. Covariates considered for inclusion in the model were identified a priori without knowledge of the outcome data based on clinical judgement and potential confounders identified in the literature.^21–24^ Backwards elimination, recursive feature elimination, and clinical relevance were used for final predictor selection. Scikit-Learn, an open source library was used for machine learning modeling and AUC.^25^ Multivariable logistic regression with standard errors was used to determine the independent association of factors and severe COVID-19. The β coefficient and adjusted odds ratio (OR) with 95% confidence intervals (CI) were obtained for each predictor. The model constant and β coefficient for each predictor were used to generate the predicted probability equation. The prediction model was assessed by the calibration plot.

The discriminative ability and performance of the model was assessed by calculating the AUC. The Hosmer–Lemeshow test was used to assess goodness-of-fit. Statistical significance was set a priori at p<LJ0.05. Python (version 3.6.6), Statsmodels (version 0.9.0, for regression), and RStudio (version 1.1.463) were used for statistical analysis. The final model’s predicted probability equation was incorporated into a mobile-friendly web-based application (www.covidmodel.org) developed using Scikit-Learn module. Despite institutional COVID-19 treatment guidelines, not all patients had uniform laboratory testing done at the time of presentation, resulting in some missing data. We classified these missing laboratory values as separate categories and tested their association with the primary outcome. Patients with missing CRP and AST values had the lowest rates of severe COVID-19 and behaved similar to patients with laboratory values within normal reference range. Therefore, these patients were combined into the same category as patients within the normal reference range and kept in their respective derivation or validation cohorts.

The Thomas Jefferson University Institutional Review Board approved this study and waived informed consent from study participants.

## Results

### Participant Characteristics

A total of 415 patients with COVID-19 were included, of whom 164 (39%) developed severe COVID-19. The median age was 66 years (IQR 54-77). The majority were male (55%) and most were Black (44%), followed by White (34%), Asian (13%), and Hispanic (8%) (which our EHR categorized under race). **Table 1** shows univariate analyses of patient demographic and clinical characteristics. Patients who developed severe COVID-19 were more likely to be older (age 70.6 vs 61.4, p<0.001), male (66.3% vs 48.0%; p<0.001), and present with dyspnea (75.5% vs 61.5%; p=0.004). They were also more likely to have a past medical history of diabetes (44.8% vs 28.6%; p=0.001), coronary artery disease (23.3% vs 14.7%; p=0.04), and/or prior stroke (20.2% vs 11.9%; p=0.03). Analyses of laboratory data upon presentation showed that severe COVID-19 was associated with higher levels of CRP, AST, and D-dimer (**Figure 1-A**). Notably, a CRP of 0-5 mg/dL was associated with a 27.5% rate of severe COVID-19 compared with 40.9% in those with a CRP of 5.1-15 mg/dL and 72.5% in those with a CRP >15 mg/dL. D-dimer 0-300 ng/mL was associated with a 26.7% rate of severe COVID-19 compared with 38.9% in those with D-dimer 300.1-1000 ng/mL and 68.8% in those with D-dimer >1000 ng/mL.

**Figure 1.**
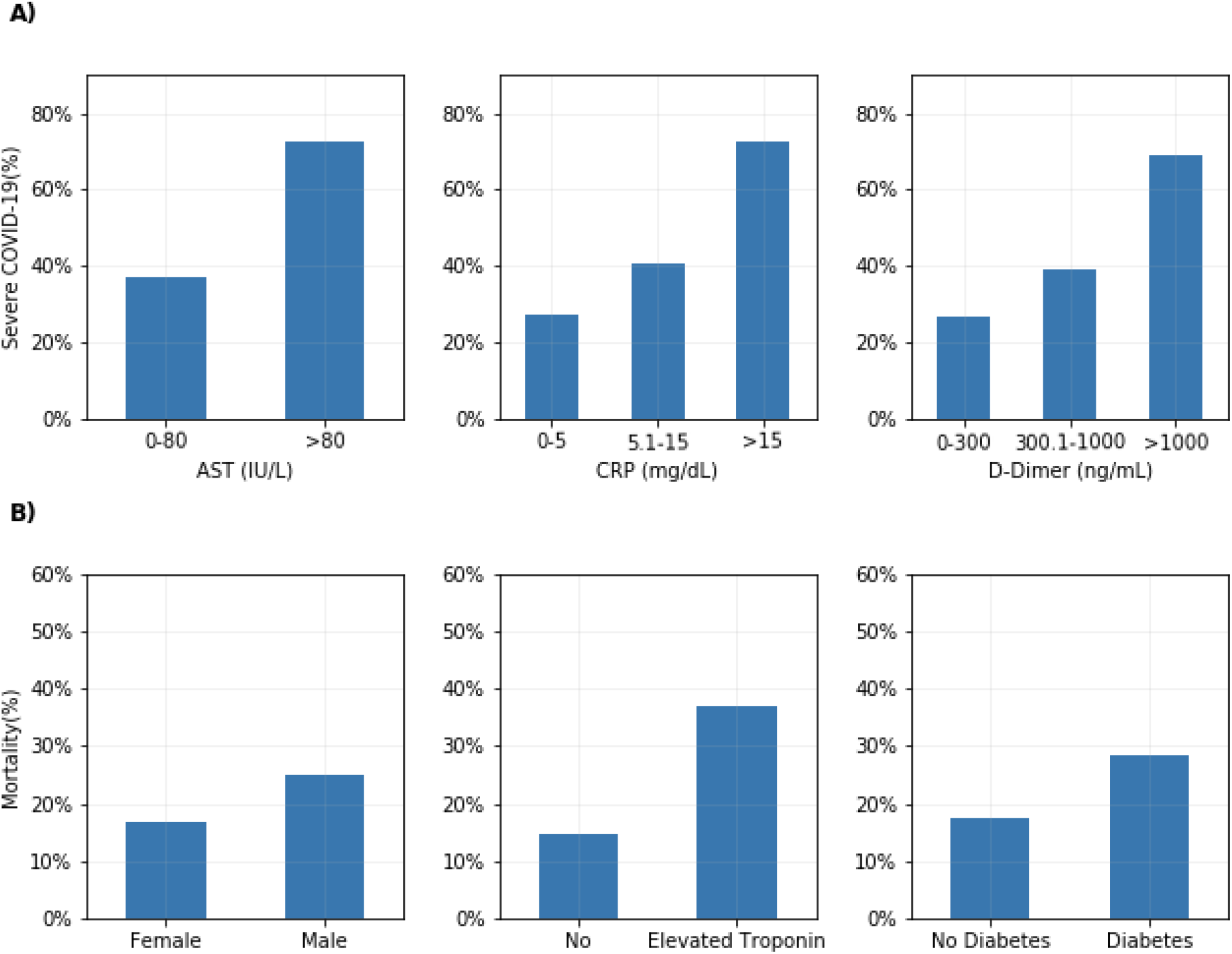
Association of selected (A) laboratory factors with severe COVID-19 and (B) patient-level factors associated with COVID-19 mortality. **Footnote:** Elevated troponin: HsTroponin T>19ng/L or Troponin I≥0.04ng/mL

**Table 1.** Demographic, clinical, and laboratory characteristics of patients with COVID-19 upon presentation, by severity.

|  | Non-Severe COVID-19<br>(n=252) | Severe COVID-19<br>(n=163) | p-value |
| --- | --- | --- | --- |
| Age, Mean (±SD) | 61.4 (±15.8) | 70.6 (±15.1) | **<0.001** |
| Male (%) | 48.0 | 66.3 | **<0.001** |
| Diabetes (%) | 28.6 | 44.8 | **0.001** |
| Body mass index | 30.6 | 30.5 | 0.57 |
| Hypertension (%) | 66.5 | 75.0 | 0.05 |
| History of cancer (%) | 10.8 | 17.3 | 0.08 |
| Coronary artery disease (%) | 14.7 | 23.3 | **0.04** |
| Congestive heart failure (%) | 13.3 | 20.4 | 0.07 |
| Stroke (%) | 11.9 | 20.2 | **0.03** |
| Chronic obstructive pulmonary disease (%) | 14.7 | 22.7 | 0.05 |
| End-stage renal disease (%) | 4.4 | 4.9 | 0.99 |
| Dyspnea (%) | 61.5 | 75.5 | **0.004** |
| Initial white blood cell count<br>($\times 10^9$/L) | 6.6 | 9.1 | **<0.001** |
| Initial hemoglobin (g/dL) | 12.7 | 12.9 | 0.34 |
| Initial sodium (mEq/L) | 136.1 | 136.9 | 0.59 |
| Initial creatinine (mg/dL) | 1.61 | 2.04 | **<0.001** |
| C-reactive protein (mg/dL) | 6.61 | 12.1 | **<0.001** |
| Aspartate aminotransferase (IU/L) | 44.0 | 88.2 | **0.002** |
| Alanine aminotransferase (IU/L) | 34.8 | 50.6 | 0.44 |
| Ferritin (ng/mL) | 742.6 | 1306.3 | **<0.001** |
| D-dimer (ng/mL) | 730.0 | 2324.7 | **<0.001** |

**Figure 1-B** shows unadjusted rates of 14-day mortality for sex, elevated troponin, and a history of diabetes. Male sex had a higher rate of mortality (24.9% vs 16.7%, p=0.06). Patients with diabetes had an increased rate of mortality (28.3% vs 17.4%, p=0.01). Elevated troponin was associated with a higher rate of mortality (37.2% vs 14.8%, p<0.001). In multivariable analysis, male sex (OR 2.44, p=0.01), diabetes (OR 2.44, p=0.01), and elevated troponin (OR 2.21, p=0.02) had significantly increased odds ratio of mortality (**Table 3**).

**Table 2.** Coefficients, adjusted odds ratio of severe COVID-19 predictors in multivariable logistic regression.

|  | **Coefficient** | **SE** | **p-value** | **Adjusted Odds ratio (95% CI)** |
| --- | --- | --- | --- | --- |
| Intercept | -5.411 | 0.865 |  |  |
| Age | 0.037 | 0.01 | <0.001 | 1.04 (1.02-1.06) |
| Sex (Male) | 1.113 | 0.297 | <0.001 | 3.04 (1.7-5.44) |
| Dyspnea | 0.671 | 0.317 | 0.034 | 1.96 (1.05-3.64) |
| Diabetes | 0.827 | 0.288 | 0.004 | 2.29 (1.3-4.02) |
| C-reactive protein (mg/dL)<br>(reference:0-5 mg/dL) |  |  |  |  |
| 5.1-15 mg/dL | 0.567 | 0.311 | 0.068 | 1.76 (0.96-3.25) |
| >15 mg/dL | 1.650 | 0.409 | <0.001 | 5.21 (2.33-11.61) |
| Aspartate aminotransferase<br>>80 (IU/L) | 1.355 | 0.46 | 0.003 | 3.88 (1.57-9.56) |
| D-dimer (ng/mL)<br>(reference: 0-300 ng/mL) |  |  |  |  |
| 300.1-1000 ng/mL | 0.031 | 0.378 | 0.935 | 1.03 (0.49-2.16) |
| >1000 ng/mL | 0.951 | 0.467 | 0.042 | 2.59 (1.04-6.47) |
| Elevated troponin<br>(HsTroponin T > 19ng/L, or<br>Troponin I ≥ 0.04ng/mL) | 0.399 | 0.302 | 0.187 | 1.49 (0.82-2.7) |

**Table 3.** Coefficients, adjusted odds ratio of COVID-19 14-day mortality predictors in multivariable logistic regression.

|  | Coefficient | SE | P value | Adjusted odds ratio |
| --- | --- | --- | --- | --- |
| Intercept | -9.7919 | 1.371 |  |  |
| Age | 0.0762 | 0.015 | <0.001 | 1.08 (1.05-1.11) |
| Sex (Male) | 0.8904 | 0.359 | 0.01 | 2.44 (1.21-4.92) |
| Dyspnea | 0.6086 | 0.384 | 0.11 | 1.84 (0.87-3.9) |
| Diabetes | 0.8925 | 0.351 | 0.01 | 2.44 (1.23-4.86) |
| C-reactive protein (mg/dL)<br>(reference:0-5) |  |  |  |  |
| 5.1-15 | 0.3967 | 0.394 | 0.31 | 1.49 (0.69-3.22) |
| >15 | 0.908 | 0.468 | 0.05 | 2.48 (0.99-6.2) |
| Aspartate aminotransferase<br>(IU/L) >80 | 1.0719 | 0.515 | 0.04 | 2.92 (1.06-8.01) |
| D-dimer (ng/mL)<br>(reference: 0-300 ng/mL) |  |  |  |  |
| 300.1-1000 | 0.8894 | 0.619 | 0.15 | 2.43 (0.72-8.18) |
| >1000 | 1.3223 | 0.678 | 0.05 | 3.75 (0.99-14.17) |
| Elevated troponin<br>(HsTroponin T > 19ng/L, or<br>Troponin I ≥ 0.04ng/mL) | 0.7913 | 0.348 | 0.02 | 2.21 (1.11-4.37) |

### Primary and secondary outcomes

The percentage of patients developing the primary outcome of severe COVID-19 was 39.3% (n=163). Analysis of individual outcomes showed that 31.6% (n=131) of patients were admitted to the ICU, 24.6% (n=102) required mechanical ventilation, and 21.2% (n=88) died within 14 days of admission. **Figure 2** shows the distribution of hospital days in which patients first required mechanical ventilation. Mean time to mechanical ventilation was 3.73 hospital days (SD 2.7). After the first two days, 55.9% (n=57) required mechanical ventilation (**Figure 2**).

**Figure 2.**
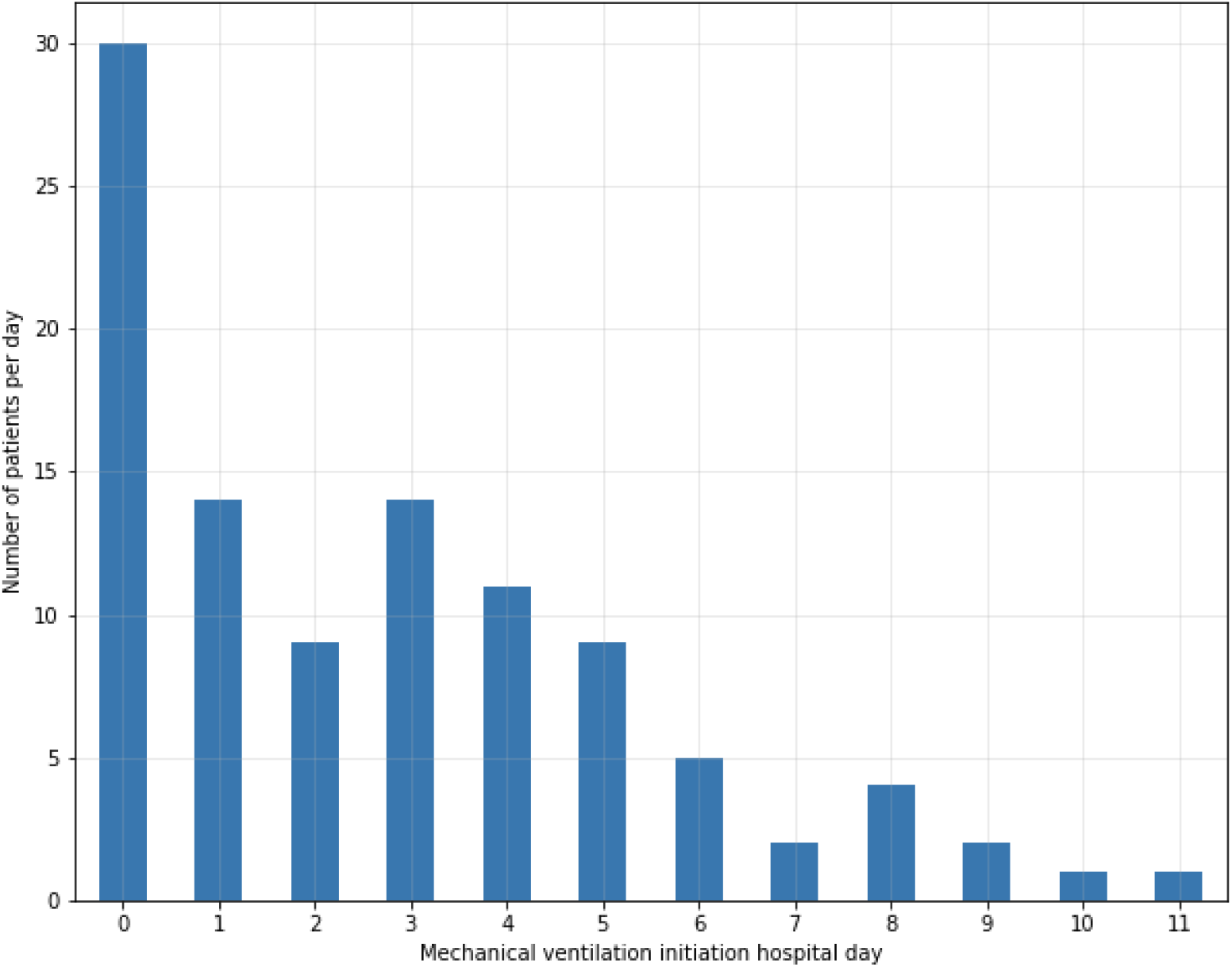
Distribution of hospital day on which patients required mechanical ventilation.

### Prediction of severe COVID-19 and validation of risk prediction model

Multivariable analyses on the training cohort demonstrated that increased age, male sex, diabetes, dyspnea, CRP > 15 mg/dL, AST > 80 IU/L, and D-dimer > 1000 ng/mL were independently associated with severe COVID-19 (**Table 2**). The strongest predictors of severe COVID-19 were AST > 80 IU/L (OR 3.88 [95% CI 1.57-9.56], p=0.003), male sex (OR 3.04 [95% CI 1.70-5.44], p<0.001), and diabetes (OR 2.29 [95% CI 1.30-4.02], p=0.004) (**Figure 3**). Variables included in the risk prediction model were age, sex, diabetes, dyspnea, CRP, AST, D-dimer, and troponin.

**Figure 3.**
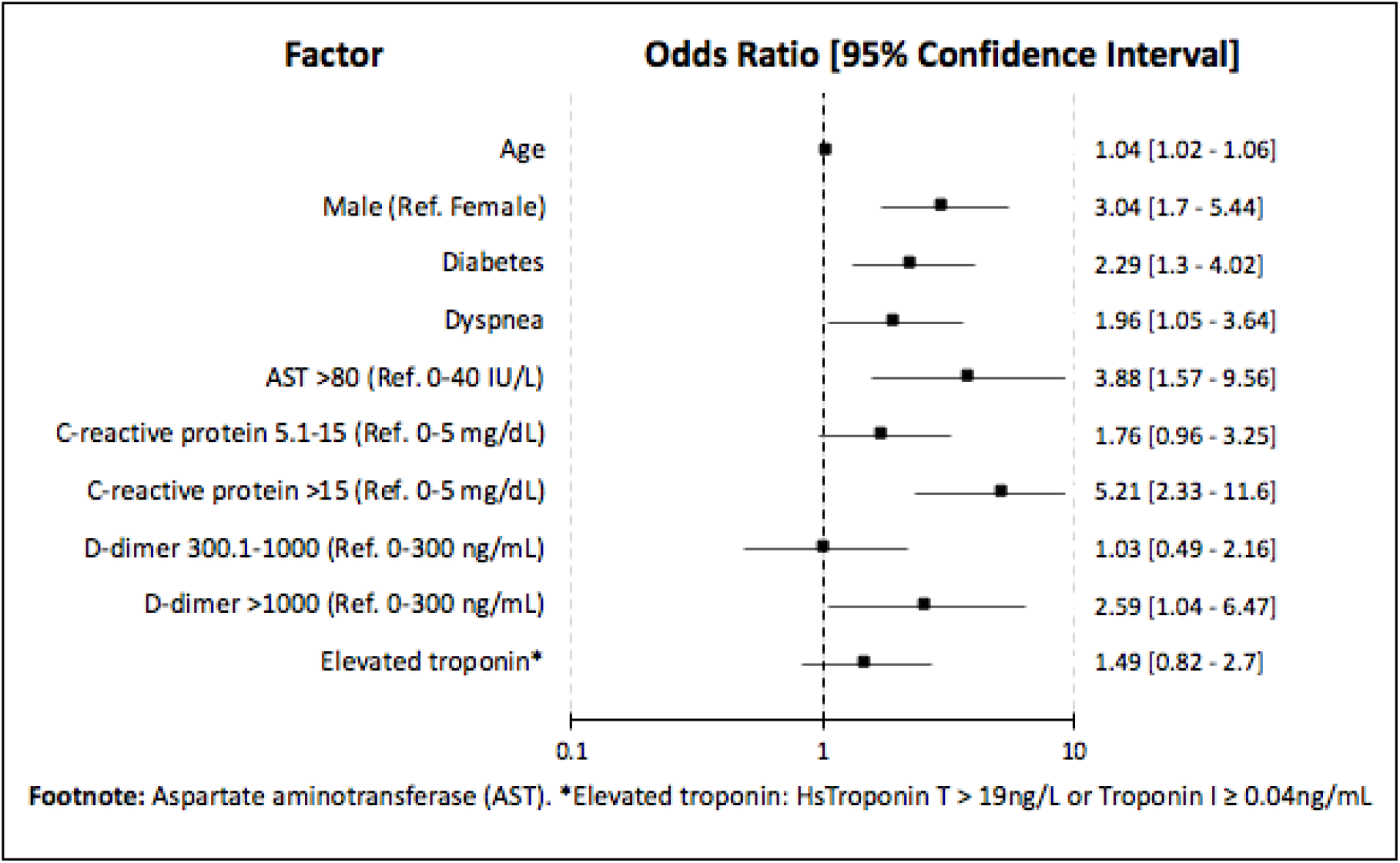
Forest plot showing the independent association of patient demographic, clinicalcharacteristic, and laboratory data with severe COVID-19 composite outcome (mortality, ICU admission, mechanical ventilation).

The model showed excellent calibration as it was well matched with the 45-degree line (**e-Figure 2**) and excellent goodness-of-fit on a Hosmer and Lemeshow test (p =0.22). This severe COVID-19 model exhibited excellent discriminative ability (AUC: derivation cohort=0.82, validation cohort=0.82). The COVID-19 14-day mortality prediction model also demonstrated excellent performance (AUC: derivation cohort=0.85, validation cohort=0.81). **e-Table 1 and 2** show adjusted odds ratio of the eight predictors used in the models including both derivation and validation cohorts combined. The model’s sensitivity was 83%, and specificity was 60% with the cutpoint probability of 23%. Sensitivity analysis after removing sex predictor showed severe COVID-19 model AUC (derivation cohort=0.79, validation cohort=0.80).

### Application of severe COVID-19 risk model

The probability of the occurrence of severe COVID-19 and COVID-19-related death can be calculated using the predicted probability equation, which is based on the model’s intercept and coefficients from **Table 2**. A mobile web-based COVID-19 risk model is shown in **e-Figure 3**.

#### Case 1

A 60-year-old female with a history of diabetes presents to the hospital with dyspnea and cough. Labs show hsTroponin T within normal limits, AST 65 IU/L, CRP 12 mg/dL, and D-dimer 280 ng/mL. Her 14-day risk of severe COVID-19 is 25.3%. Her 14-day risk of death is 4.2%.

#### Case 2

A 65-year-old male with no history of diabetes presents with dyspnea. Lab show hsTroponin T 35 ng/L, AST 70 IU/L, CRP 8 mg/dL, D-dimer 700 ng/mL. His 14-day risk of severe COVID-19 is 44.8%. His 14-day risk of death is 22.6%.

## DISCUSSION

COVID-19 associated morbidity, mortality, and stress on the healthcare system is expected to continue. To address the on-going pandemic, the addition of reliable and easy-to-use models of deterioration to severe COVID-19 will better allow clinicians and systems to make improved, evidenced-based patient care decisions. This study developed and internally validated a web-based model to predict 14-day risk of progression to severe COVID-19 using a cohort of 415 diverse U.S. patients hospitalized at a large academic medical center and four community hospitals. Compared to other large case studies in the U.S., patients in our study have similar demographics, clinical characteristics, and mortality rates.^9,26^

There have been several studies from China, Italy, and the U.S. that have identified characteristics of patients who have poor outcomes from COVID-19 based on retrospective analyses.^7,9,27–29^ Similar to Zhou et al., we identified an increased risk of severe COVID-19 with older age.^13^ An analysis of 1,150 adults in New York City hospitals indicated that chronic pulmonary disease, followed by cardiovascular disease, older age, higher concentrations of interleukin-6, and D-dimer at admission were the strongest predictors of mortality with Black and Hispanic patients presenting later in the disease course compared to White patients.^30^ These findings are similar to the clinical predictors used in our study, in which an elevated CRP and D-dimer are suggestive of a profound inflammatory state. In agreement with data from a multicenter observational study, we found increased odds of death with male sex and history of diabetes.^31^ Similarly, positive troponin was found to be associated with increased odds of death (OR=2.21, p=0.02) which is also consistent with prior studies. ^21,32^ For this reason and practical purposes, troponin was included in both the severe COVID-19 and mortality models.

Assessment of time to mechanical ventilation revealed that 55.9% were mechanically ventilated after two days of hospitalization. Currently, there are no easy-to-use and effective U.S.-patient-population-based methods to identify these high risk patients whose clinical picture at the time of presentation does not suggest severe COVID-19. This point becomes particularly relevant if one considers that a quarter (24.5%) of patients who developed severe COVID-19 did not have dyspnea on presentation. Existing risk calculators in the literature are derived from clinical and laboratory data from China that have not been externally validated in a U.S. population.^17,33–36^ The present model addresses this knowledge gap by providing a validated prediction model that quantifies the risk for severe disease and mortality in COVID-19 derived from a U.S. patient population and by incorporating it into a mobile-friendly model that can be easily accessed by anyone with internet access globally.^36^

The model’s potential applications are numerous. It can be leveraged to inform more specific inclusion/exclusion criteria for COVID-19 clinical trials. Clinicians may use this model to make informed risk-benefit decisions regarding Emergency-Use-Authorization, off-label, and other potential therapies. The model may help hospitals and/or hospital systems to predict imminent ICU bed and mechanical ventilator needs. Last, this model may aid emergency medicine and non-ICU hospital clinicians to more effectively and safely triage patients to an appropriate level of care and to make admission, discharge, and post-discharge monitoring and follow-up decisions.

This study has several strengths. First, the study participants represent a diverse self-reported race of patients residing in three states. Second, this population represents a cohort of inpatients that reflect common U.S. comorbid conditions which will allow wide applicability for clinical use in the U.S. and other countries with similar demographics and comorbidity types and rates. Third, this study is one of the first prediction models in the U.S.. Fourth, the data used for the model development is derived from multiple hospital types including a large academic center and several community hospitals thereby reducing the likelihood that observed outcomes are confounded by a single institution’s unique treatment approach. Last, the model is available for real-time clinical use via a web-based calculator.

This study is not without limitations. First, the data was extracted retrospectively, relied on EHR provider documentation, and was limited to variables contained in the EHR. To help ensure data validity, the variables and outcomes of interest were extracted by physician-investigators and validated by an independent researcher. Second, the sample size is relatively small compared to larger studies from China. These models had excellent performance during the internal validation process, therefore, we chose to prioritize the dissemination given the urgent need of prediction models tailored specifically to the U.S. to care for patients suffering from COVID-19. Third, given the rapidly changing “standard of care” for COVID-19 and institutional efforts to educate clinicians in near real-time, there was likely significant practice variation both within each hospital and between hospitals between March 1, 2020 and April 30, 2020 that might affect outcomes. Nonetheless, we have provided a mobile-friendly model for prediction of severe COVID-19 upon presentation.

## Conclusion

In conclusion, this study presents an internally-validated prediction model for progression to severe COVID-19 and mortality in hospitalized patients that can be used in real-time at the bedside.

## Data Availability

Data is available

## Acknowledgments

The authors would like to acknowledge and thank Chris J. Li, Conor G. Bradley, Christa M. Smaltz, MD, Crystal Y. Lee, MPH, David B. Ney, Danielle M. Fitzpatrick, MD, Joseph W. Schaefer, Kashyap Chauhan, MD, Margaret V. Szot, MD, and Shuji Mitsuhashi, MD for their extensive and selfless contribution to this project and the patients it seeks to serve.

## Author contributions

S.H.W. is the guarantor of the paper. S.H.W, A.J.R, A.A.K, D,R.C, L.L.A, D.M.C, M.B. had full access to all the data. S.H.W. built the models for a website. S.H.W, A.J.R, A.A.K, D.R.C., L.L.A.,M.B. were involved in study design. S.H.W, A.J.R, A.A.K, D.R.C.,L.L.A, D.M.C, C.M.V, J.M.R were involved in data collection and validation. All authors were involved in interpretation of data and drafting of the manuscript.

### Funding/Support

*no funding*

## FIGURES AND TABLES LEGENDS

**Figure 1-A**. Association of AST, C-reactive protein, or D-Dimer with severe COVID-19, defined as ICU admission, mechanical ventilation, or death.

**Abbreviations:** Aspartate Aminotransferase (AST); C-reactive protein (CRP)

**Figure 1-B**. Unadjusted 14-day mortality rate and sex, elevated troponin and history of diabetes.

**Footnote:** Elevated troponin: HsTroponin T>19ng/L or Troponin I≥0.04ng/mL

**Figure 2**. Distribution of hospital day in which patients required mechanical ventilation.

**Figure 3**. Forest plot showing the independent association of patient demographic, clinical characteristics, and laboratory data with the severe COVID-19 composite outcome (mortality, ICU admission, mechanical ventilation).

**Footnote:** *Elevated troponin: HsTroponin T>19ng/L or Troponin I≥0.04ng/mL

**Table 1**. Demographic, clinical, and laboratory characteristics of patients with COVID-19 upon presentation, by severity.

**Table 2**. Coefficients, adjusted odds ratio of severe COVID-19 predictors in multivariable logistic regression.

**Table 3**. Coefficients, adjusted odds ratio of COVID-19 14-day mortality predictors in multivariable logistic regression.

**e-Figure 1**. Theoretical phases of COVID-19 disease course.

**e-Figure 2**. Calibration plot of the severe COVID-19 prediction model.

**e-Figure 3**. Mobile web-based severe COVID-19 risk model.

**e-Table 1**. Adjusted odds ratio of severe COVID-19 predictors in multivariable logistic regression. All patients (n=415)

**e-Table 2**. Adjusted odds ratio of 14-day mortality predictors in multivariable logistic regression. All patients (n=415)

